# Impact of Covid-19 on Bangladeshi university students mental health: ML and DL analysis

**DOI:** 10.1101/2024.05.17.24307476

**Authors:** Md Monir Ahammod Bin Atique, Md Ilias Bappi, Kyungbeak Kim, Kwanghoon Choi, Md Martuza Ahamad, Khondaker Masfiq Reza

**Affiliations:** Department of Computer Science and Engineering, Uttara University, Dhaka, Bangladesh.; Department of Artificial Intelligence Convergence, Chonnam National University, Gwangju, South Korea.; Department of Computer Science, Marquette University, Milwaukee, United States.; Department of Computer Science, Colorado State University, Colorado, United States.

**Keywords:** Covid-19, Coronavirus, Bangladesh, Machine Learning, Deep Learning, Mental Health, Pandemic, University students

## Abstract

The Covid-19 outbreak has adversely influenced university students across the world both physically and psychologically. The psychological struggle faced by students, is effected by various factors, including disruptions to daily routines and academic activities, increased reliance on smartphones and the internet, limited social interaction, and confinement to their homes. These impediments reflect a broader issue of imbalance in cognitive health status among them during the pandemic. In Bangladesh, despite having the necessary population to study, understanding the impact of Covid-19 on the mental health status of university students has been limited. Hence, it is imperative to diagnose mental health issues and deal with the underlying reasons in order to enhance students’ psychological well-being, which leads to academic achievement. Nowadays, Artificial Intelligence (AI) based prediction models have the potential to play a crucial role in predicting mental state early. The purpose of the study is to explore the following effects of the pandemic on the mental health of Bangladeshi university students using Machine Learning (ML) and Deep Learning (DL) techniques. A reliable AI prediction system requires real-world data, that was collected by a survey through a Google form (online questionnaires) among 400 students of 16 universities, and the respondents were 253. In this paper, after data preprocessing, ten widely known ML and four DL models were developed to automatically and accurately predict mental well-being during or after the Covid-19 circumstance. According to our findings, the Random Forest (RF) algorithm and Siamese Neural Networks (SNNs) outperformed other models in terms of accuracy (86% and 75%). Additionally, Chi-Square test was conducted, which revealed the five most common and significant predictors (“Stable family income”, “Disruption of daily life”, “Own income”, “Sleep status”, and “Fear of getting infected with Covid-19”) of psychological health conditions. Overall, this work could assist university administrations, government agencies, and health specialists in taking appropriate measures to understand and maintain students’ mental health. This research also suggests proper monitoring, government support, and social awareness during and after the worldwide epidemic to keep an excellent mental health state of university students.

## 1 Introduction

Mental health refers to general psychological condition of any individual. It is also defined as general ability to deal with people, stresses as well as one’s contribution in the community.[1] Several reasons have been identified through numerous studies [2–4] regarding the importance of maintaining good mental health conditions in daily life. One of the most significant reasons is that every person has a direct link between his or her physical and mental health. According to medical science, a patient with a chronic disease is said to have a close connection to the possibilities of developing depression [5]. Studies also show correlations among mental health well-being and social relationships within families and communities [6]. Therefore, it is obvious that mental health status has a direct impact on individual personnel’s and social development. A survey shows that individuals with balanced mental health are more keen on personal advancement through appropriate planning and intentional engagement [7]. However, healthy psychological state is affected when a global pandemic arises.

In 2019, people were first introduced to the Novel Coronavirus (Covid-19) disease, which was derived from SARS-Cov-2 (Severe Acute Respiratory Syndrome Coronavirus-2). Before December 31, 2019, the globe had never faced such a disease. It contributed to lung injury, cardiac injury, and even death. The World Health Organization (WHO) declared Covid-19 as a global pandemic on March 11, 2020. During the mass contagion, social isolation was implemented by the governments of many countries. This outbreak had reached Bangladesh as well. As of 2020, Bangladesh was the ninth most densely populated country and territory in the world [8]. Considering this huge population, the Bangladeshi government also implemented lockdowns all over the country to protect against the devastating scenario. As a result, many young adults have had to face a new kind of environment where going out of the home and maintaining social relationships have become challenging or impossible, leading to a new circumstance called “social isolation”, as mentioned earlier. This situation had a great impact on people’s mental health, especially on those who already had a mental illness before the health disaster. Social isolation may lead to loneliness which can make a person more vulnerable to ill mental conditions like depressive symptomatically, higher suicide attempts, and completed suicide [9]. This may also lead to dementia -which refer to not being able to think, remember, make decisions for everyday activity [9]. As a strong linkage between mental health and social isolation, it is found that the longer the isolation may take place, people become more mentally sick and more vulnerable to depression and anxiety [10]. Young people are the ones who suffer the most from this isolation.

University students are similar to other young adults in acknowledging the importance of addressing mental health needs. In other words, they are not unique in this regard; they share the same awareness of the importance of mental health as other young generation. This infectious disease crisis severely affected their academic and social activities. During this time, students stayed home and faced several problems, such as loneliness, career concerns, and financial issues. They can’t meet with their friends and relatives physically, which affects many aspects of their mental health. Most of the time, they are isolated, and sometimes they feel negative thoughts, which sometimes cause depression and anxiety. Many university students had medium to high levels of stress and suicidal thoughts at that time. Research found that students with poor mental conditions have anxiety and depression, which in some cases went as high as clinical cut-offs [11, 12]. This type of psychological degradation is less visible except during the viral outbreak.

Over the decades, there has been increasing worldwide recognition of the adverse effects of stress, anxiety, and depression on individuals, institutions, and communities [13]. In that ongoing context in recent years, Covid-19 has imposed a negative impact on mental well-being, especially on university students. Therefore, a group of studies has been conducted globally to figure out the efficient method that can aid the respected authority in identifying whose mental state is poor or better including severe levels of anxiety and depression symptoms. Many previous works have been taken into account before moving forward with this study. Most of the studies [14–22] focused on physiological and statistical techniques for exploring the factors such as gender, education status, etc which have a relation of being mentally sick. However, most of the above studies did not concentrate on AI-driven methods such as ML, DL, and Data Mining. Similar to the other fields, AI technologies can be used in the field of mental health for quite some time. Various ML and DL techniques [23–37] have been utilized to analyze the mental health condition of any individual with descent accuracy. Different mainstream algorithms such as Decision Tree (DT), RF, Support Vector Machine (SVM), Naïve Bayes (NB), Logistic Regression (LR), Extreme Gradient Boosting (XGB), Gradient Boosting Classifier (GBC) and Artificial Neural Networks (ANNs) including Convolutional Neural Network (CNNs) and Recurrent Neural Networks (RNNs) have been developed several times for the prediction. Nevertheless, very few studies have been conducted exploring the university (undergraduate, graduate, and Ph.D.) students data. Although Covid-19 has a considerable influence on the psychological well-being of students at higher education institutions, there is comparatively less research on predicting mental health states. Particularly in highly populated countries like Bangladesh, there exists a research limitation in developing practical and reliable studies on university students. While there were several studies [13, 38] in Bangladesh, there’s still plenty of room to improve both the statistical analysis of features and the application of DL techniques. Generally, research on mental health status is not given the same level of attention as their physical health in Bangladesh. Another limitation of the existing research is not using real-world datasets, and that is why there is enough scope for improvement in the data collection methods.

The main goal of our study is to build an AI-driven system including proper analysis of features where we utilized our primary dataset which is based on real feedback on Covid-19’s impact from Bangladeshi university students. During the experiment, we collected data through an online questionnaire and trained not only ten well-known ML models such as RF, CatBoost, XGB, GBC, SVM, DT, K-Nearest Neighbors (K-NN), Light Gradient Boosting Machine (LGBM), AdaBoost, and Multinomial Naive Bayes (MNB) but also four DL techniques including CNNs, RNNs, Hybrid Neural Network, and SNNs. These models were then assessed on testing subset how well they predicted the students’ well-being status (Poor or Better). Each model was evaluated against performance metrics. Finally based on this evaluation, best-performing model (RF and SNN) was determined. Additionally, we also performed Chi-Square test as a statistical associativity of features which contributed to figuring out the most associative attributes influenced on mental well-being. Through this test, we found five highly significant factors (“Stable family income”, “Disruption of daily life”, “Own income”, “Sleep status”, and “Fear of getting infected”) that greatly influenced psychological health conditions during this epidemic of Bangladeshi higher education students.

The following are primary contributions of our proposed research work in the context of mental health prediction:

- A real-world dataset was collected through an online survey that have a huge impact on the student’s mental health status.
- The descriptive data analysis was applied to obtain valuable insights from the dataset. The factors that affect mental health was examined thoroughly.
- Performed a statistical test of the features to understand how these features contribute to the level of mental health status.
- To accurately predict student’s mental health labels (Poor or Better) using AI-driven classification models.
- To determine the adverse impact of social isolation on university students mental and psychological well-being during the Covid-19 outbreak.
- Our findings can assist government agencies, university authorities, and healthcare practitioners to ensure university students’ mental fitness and provide them with practical solutions to maintain a healthy educational experience.

The rest of the paper is organized as follows: Related Works describes the similar works. Then, Research Methodology discusses the methodology and presents a brief description of the dataset, descriptive data analysis of the features, preprocessing tasks, feature selection and ranking, proposed models, and evaluation criteria of the applied models. Experimental Results Analysis shows Chi-Square test, ML, and DL result analysis. Discussion presents the general discussions of the whole study, limitations and strengths. At last, Conclusion demonstrates the concluding statements of this study, and mentions the direction of future research.

## 2 Related Works

Mental health issues encompass a broad range of conditions that affect thoughts, emotions, and behaviors, thereby impacting an individual’s overall well-being. These conditions may create anxiety, depression, stress, or other forms of psychological imbalance that affect daily life. Covid-19 significantly impacted mental health during the pandemic, prompting many studies to be conducted both during and after this period. Below, we discuss some relevant research works pertaining to our study.

### Mental Health Status, Anxiety, and Depression Levels

Previously related works [14–16] found many factors such as age, gender, family type, marital status, education, and employment status that directly contribute to mental health issues among the students doing higher studies. Many studies conducted investigation ground on statistical surveys in the United States and China [17–20] reveals the adverse impact of Covid-19 prevalence on mental health especially amongst college or university students. Their findings found the existence of poor mental health among most university students during the pandemic period. In China, 24.9% Chinese university students faced anxiety situations in that time [17]. Another research conducted in the United States [18] colleges during the Covid-19 pandemic revealed that 24.5% of 1881 students experienced depression (PHQ-9), while 20.7% reported anxiety (GAD-7). Additionally, the experiment highlighted the adverse effects of remote learning on personal relationships and identified poor sleep quality as a contributing factor to higher risks of depression and anxiety. Similar to other countries around the world, Covid-19 has had a devastating effect on Bangladesh’s education sector. On March 17, 2020, the government of Bangladesh closed all schools, colleges, and institutions in the country due to the presence of eight verified cases to avoid these devastating diseases. According to statistics [39]. in Bangladesh, approximately 1 million university students are doing their bachelor (undergraduate), graduate, and PhD studies at 53 public, 107 private, and 3 international universities. Some researchers in Bangladesh did a few experiments on the impact of Covid-19 on university students’ mental well-being and found mild to severe-level depression symptoms. One study [21] found severe anxiety symptoms amongst students who are living with families in urban areas. Moreover, the author reported that nearly 88% of students faced mild to severe anxiety symptoms throughout the pandemic. Another survey [22] showed that approximately 69% and 47% of students reported mild to severe levels of psychological impact and mild to extremely severe levels of depression, respectively due to the pandemic.

### Using ML and DL Classifier on Mental Health Prediction

Since the outbreak, epidemiological data in Bangladesh has revealed that mental health issues due to the Covid-19 and mass isolation are prominent [24]. Early and after the pandemic, one contributing factor to these mental health implications for Bangladeshis was found to be fear [25]. Diverse studies have reiterated that Covid-19 related worries and fears for Bangladeshi Diverse studies have reiterated that Covid-19-related worries and fears among Bangladeshi individuals are associated with increased mental health issues. Thus, collectively, these findings shed light on how the pandemic can affect the mental and psychological well-being of university students in Bangladesh.

In parallel, many researchers have investigated different factors affecting students’ psychological well-being during the pandemic. Cross-national research [26] and surveys [38] conducted among university students in Bangladesh. Negative perceptions and poor mental health were found to be linked during the Covid-19 outbreak in that region [40]. During the outbreak in April of 2020, a study examined how Bangladeshi university students’ mental health, anxiety, and depression levels changed as the eruption advanced [27]. University students, according to their findings, had a significant proportion of anxiety and depression symptoms and moderate to poor mental health ratings, based on the cut scores for their individual measurements. Early in the prevalence, 65.9% of Bangladeshi medical students reported low levels of worry or depression [28], and 3.3% of individuals exhibited significant symptoms of depression. Furthermore, the use of social media as a coping mechanism during this worldwide pandemic has been observed [37], with distinct gender differences in coping strategies noted.

In the realm of applying ML on mental health studies, Reshmy Krishnan et al. worked on engineering and medical students data during Covid-19 using the Hospital Anxiety and Depression Scale (HADS) assessment [29]. They conducted an online survey using a Google form which contains questionnaire, reaching 120 medical and 152 engineering students respondents. They applied different ML algorithms like SVM, which is a robust handling method of health related data, and J48 DT. The SVM performed exceptionally well in their investigation. However, the authors only used an online survey through social media, and they couldn’t take face-to-face interviews with the students. Similarly, Hanif Abdul Rahman et al. [30] explored a dataset of 17 Southeast Asian university students. Highlighting RF and adaptive boosting as optimal algorithms, the research identifies key predictors such as “sports activities”, “body mass index”, “GPA”, “sedentary hours”, and “age”. The dataset was gathered through a cross-sectional survey methodology, and it does not allow for the establishment of causal relationships. Even though the accuracy is satisfactory but still room for improvement in data collection. Mostafa Rezapour and Lucas Hansen [31] applied several ML models and statistical techniques such as Synthetic Minority Oversampling, and Chi-Square test to analyze the mental health condition based on Covid-19 mental health data. They focused on a dataset from a mental health survey conducted during the Covid-19 isolation period among front-line workers. Roles such as nurses, emergency room staff, and surgeons had to undergo unique situations like high pressure, long hours of duty, the possibility of trauma, and emotional strain, which contributed to mental health challenges. However, this type of research can be applied to the mental health conditions of university students as well.

Every day, around 2.5 quintillion bytes of data are created on social media. Examining this vast amount of data can be a valuable way to assess the impact of Covid-19 on mental health well-being [32]. The authors, Choudhury et al. [33] collected a data set of Twitter users who are diagnosed with depression by crowd-sourcing and developed ML models for predicting the risk of depression. They also used “Social engagement”, “Prescribed medicines”, “Ego network”, and “Linguistic style” of the user as features in their paper. To avoid overfitting, they used Principal Component Analysis (PCA) and used a SVM classifier with the radial basis function (rbf) kernel for depression prediction. Tianhua Chen [34] investigated the mental health status of UK university students during the plague. This analysis aims to create a framework using a method called feature permutation importance. The goal was to make different ML methods work together, even those that can’t directly tell how important each factor is. This way, it can look at how different factors affect students’ mental well-being from various angles, going beyond the usual focus on regression. The dataset used this approach in an online survey at a UK university and found that factors like “Past medical history”, “Well-being history”, and “Facing adversity” are significant for mental wellbeing. Additionally, keeping good relationships through regular communication with family and friends, along with regular exercise, generally contributes to better mental well-being.

Few studies closely support our work. Among them, Madhuri Mahalingam et al. [35] presented a investigation to forecast anxiety levels of the university students campuses in Lebanon. Using ML based techniques they experimented on student survey items, such as demographics and self-rated health, to predict anxiety symptoms among the sample of 329 respondents. The best-performing model, Multi-Layer Perceptron (MLP) obtained the highest AUC score, and the most highly ranked feature to predict anxiety was determined to be self-rated health. They collected the dataset from one public and one private Lebanese university. However, this research could be improved by adding a wide range of university students’ mental well-being data. Besides, Ishrak Jahan Ratul et al. [13] proposed several supervised ML algorithms to examine university students’ perceived social and psychological stress among 444 university students real-world data which is also a closely related study to our work. They used some tuning and statistical techniques to get better results such as PCA, Chi-Square test, Grid Search Cross-Validation (GSCV), and Genetic Algorithm. Results showed that 11.26% of individuals had high social stress, while 24.10% had extremely high psychological stress. The MLP model showed the highest accuracy when combined with PCA and GSCV. The accuracy could be improved considering large data samples and this experiment also could be extended for predicting the social and psychological stress of university students after the pandemic. In additional research, El-Sayed Atlam et al. [36] utilized statistical and ML methods to investigate the impact of the Covid-19 pandemic on the psychological well-being of university students in three Arab countries: Egypt, Jordan, and Saudi Arabia. A questionnaire comprising demographic details and five primary dimensions - digital device usage, sleep patterns, social interactions, emotional state, and academic performance was distributed among students. Analysis of 1766 responses revealed a clear link between students’ psychological health and online education during the pandemic. However, due to the small sample size, the dataset may be biased, limiting the effectiveness of the ML model’s performance.

Changing from general ML approaches, deep learning (DL) has also shown promising results in mental health prediction. Banna et al. [32] proposed a hybrid model using LSTM and CNN to detect depressive tweets related to Covid-19. Similarly, Du et al. [41] developed a DL-based mental health monitoring scheme (DL-MHMS) using CNN on EEG data to classify students’ mental states. Almeqren et al. [42] introduced a GRU-based model to predict anxiety levels during the pandemic using Arabic tweets. Despite growing interest, there remains a noticeable research gap in applying DL techniques to assess mental health among university students. Thus, our study addresses this gap using DL models well-suited for handling categorical data with structural patterns.

Overall, studies across various countries highlight the pandemic’s significant psychological impact on students. While DL and ML offer strong predictive capabilities, further interdisciplinary research is needed to develop effective mental health support tools.

## 3 Research Methodology

This section covered a detailed explanation of how the proposed methodology has been carried out. Firstly, raw data is collected from university students through online survey and then analyzed the data descriptively to produce meaningful insights. After analyzing the raw data, we moved on to the next process, which was the data-preprocessing step. Following data pre-processing, we performed “Feature Selection and Ranking” process. Then we developed ML and DL models to predict the mental health status of the students. Finally, all the models are assessed using performance evaluation metrics. The whole research methodology is depicted in Fig 1.

**Fig. 1:**
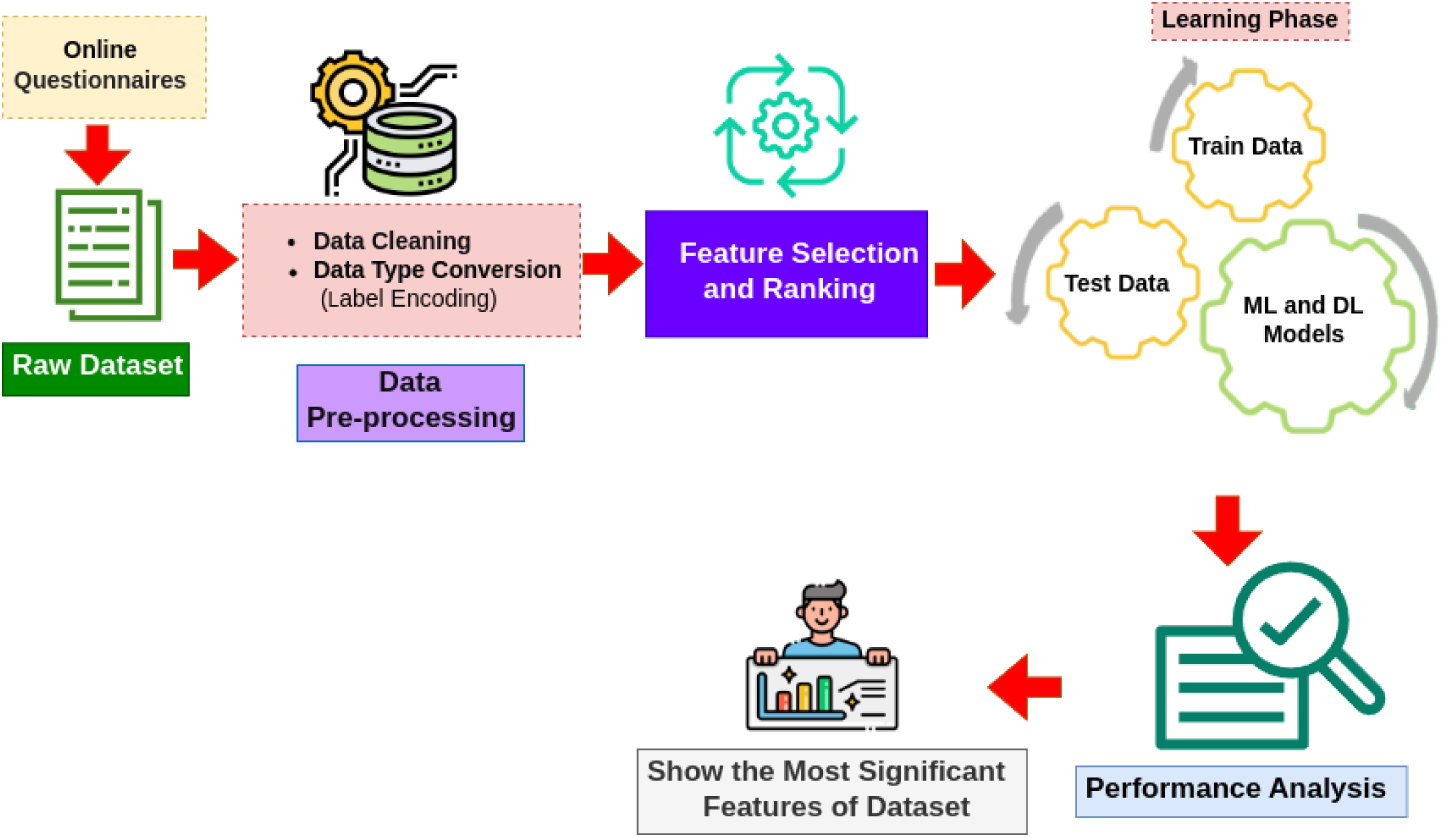
Proposed methodology.

### Dataset Description

This research focuses on university students between the ages of 19 and 30, who are the primary contributors to the dataset. From December 4, 2021, to December 13, 2021, the survey was conducted in 16 renowned universities of Bangladesh via a Google Form, collecting responses from 400 students. The form included 15 distinct questions, and a total of 253 responses were received, resulting in a 63% response rate. Among the respondents, 213 were male students and 40 were female students. A summary of the dataset is presented in Table 1.

**Table 1:**
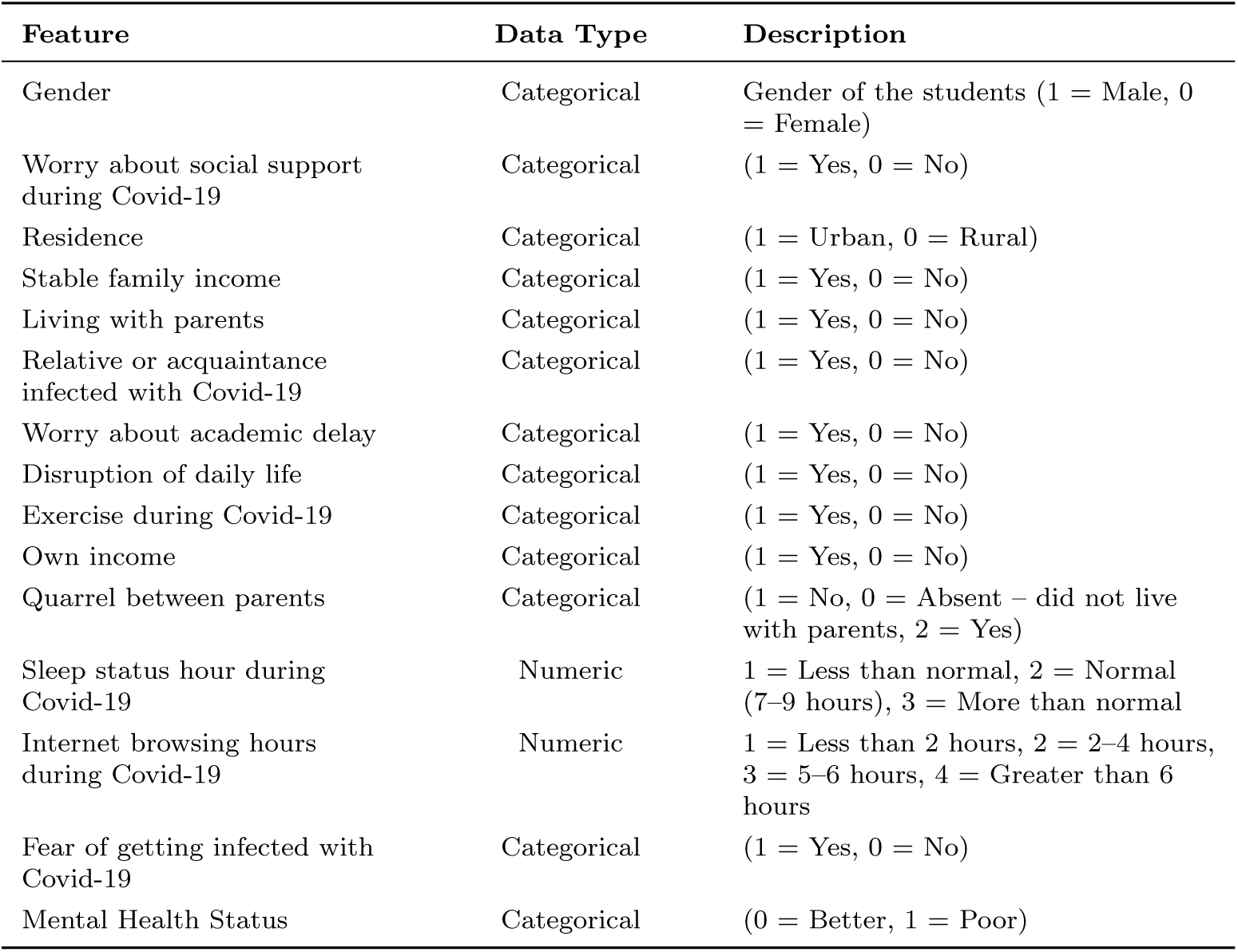
Dataset features with description.

This dataset has the following characteristics:

- This primary dataset is also considered a standard dataset and is used as a binary classification problem.
- The output column consists of two levels: *Better* and *Poor*. *Better* refers to students

who have a stable mental health state, whereas *Poor* includes students experiencing low mental well-being.

- Some key features of the dataset include *Gender*, *Stable family income*, *Worry about academic delay*, among others.
- The dataset contains 253 student entries with 15 attributes.
- Among the 253 students, 213 are male and 40 are female.

### Descriptive Data Analysis

The research revealed that fourteen features in total influence mental health status significantly. To present a proper idea of those features and how these features correlate with the label “Mental Health Status,” the statistical graph is depicted in Fig 2A, Fig 2B, and Fig 2C. Besides, a brief description with intuitive analysis is enlisted below for better understating. In this comparison, we examine the “Poor” mental health status between the “Yes” and “No” categories of features, including “Disruption of daily life”, “Worry about academic delay”, “Exercise during Covid-19”, “Worry about social support during Covid-19”, “Stable family income”, “Fear of getting infected with Covid-19”, and “Own income”.

**Fig. 2:**
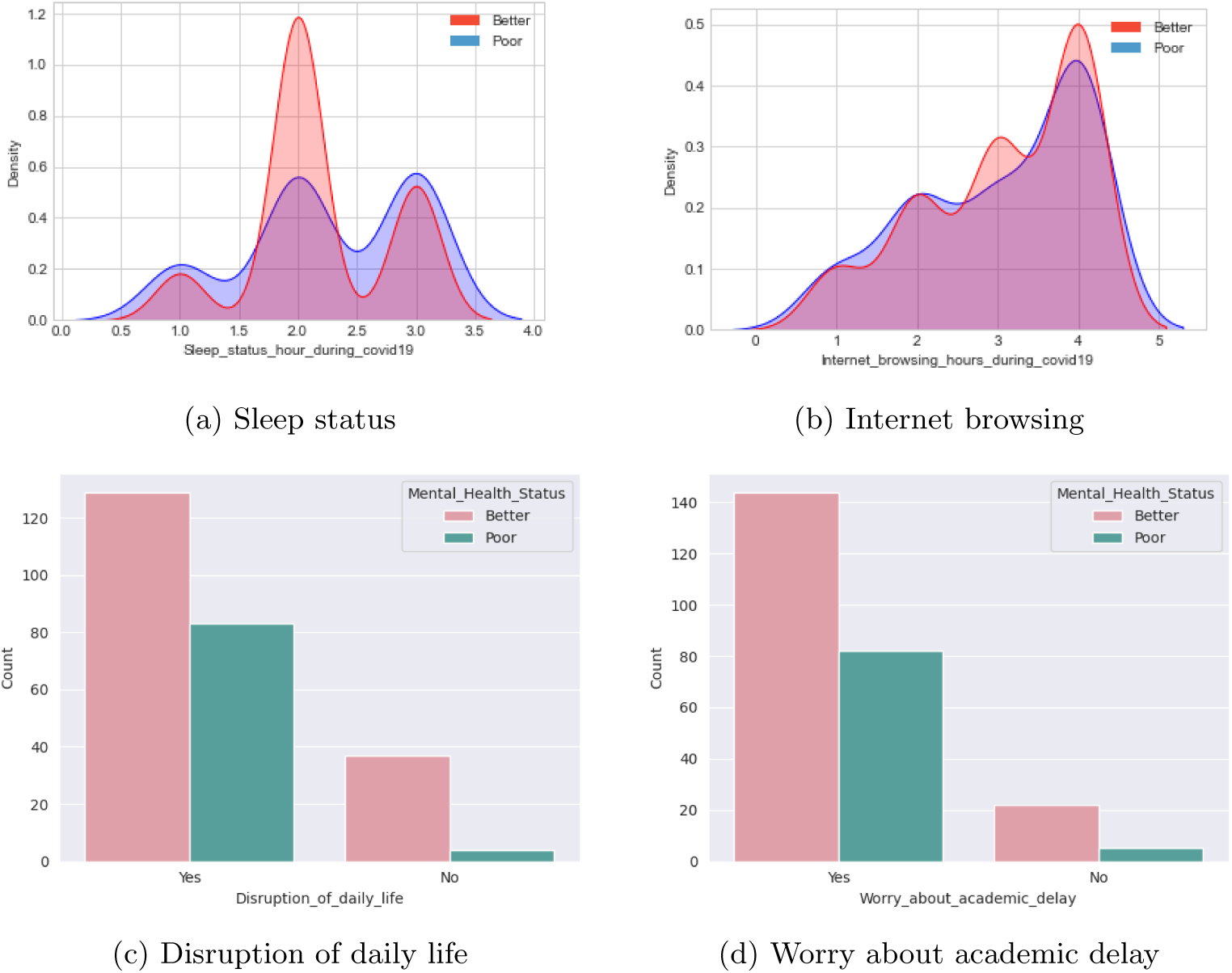
(A) Mental health status w.r.t different features.

#### Sleep status hours during Covid-19

In our analysis, we discovered that if any university student had a sleep status (hour) in the period of Covid-19 between 1.5 and 2.5, then his or her mental health status was better. The value range indicates normal (7-9 hours) or slightly less than normal (5-7 hours) sleeping hours in a day.

#### Internet browsing hours during Covid-19

Looking at the internet browsing hours of a university student, we found that if it is greater than 3 (means greater than 6 hours), then mental health status is inferior. In our exploration, we identified certain circumstances in which a student’s mental health state improves through the relaxation of their mind using the internet.

#### Disruption of daily life

Mental health status is considerably poor when disruption of life is rated “Yes”. More number of students estimated “Better” mental health state, despite the “Disruption of daily life,” features marked as “Yes”. There could be other reasons for this different scenario.

#### Worry about the academic delay

There are a considerable number of students whose mental health status is “Poor”. The reason behind this is that they are worried about academic delays. Even though a high number of students rated their mental health status as “Yes” while considering the feature “Worry about the academic delay”. There might be other features that are responsible for rating their mental health status as “Better”.

#### Quarrel between parents

If there are no quarrels between parents, then the mental health status of a student is better compared to other situations. Here, “Absent” refers to the parents’ status being late.

#### Exercise during Covid-19

Students’ mental health is relatively poor if they do not exercise during the Covid-19 epidemic.

#### Residence

Students who are in rural areas their mental health status is much better than students in urban areas.

#### Relative or acquaintance infected with Covid-19

If a student’s family or friend is not infected with Covid-19, his or her mental health will be slightly better.

#### Gender

There is a significant improvement in mental health status among males compared to females.

#### Worry about social support during Covid-19

In our research, we discovered that more students experience poor mental health conditions when they are concerned about social support.

#### Stable family income

We have seen that, when there is no stable family income, students’ mental health status tends to be poorer.

#### Living with parents

In our research, it shows that people who live with their parents act to have better mental health status than those who don’t.

#### Fear of getting infected with Covid-19

If there is a history of Covid-19 infection in the student’s family or network of friends, and there’s a fear of getting infected, then his or her mental health tends to be poorer due to the fear of infection.

#### Own income

The mental health condition of those who do not have their own income is not better compared to those who do.

### Data Preprocessing

Data preprocessing is one of the most important phases in ML and DL task, but it is difficult and time-consuming. In our experiment, we followed the preprocessing steps which is described below:

– Handling missing values: The dataset utilized in this investigation contains no missing values. Therefore, we proceed without focusing on this step.
– Data type conversion: There are 13 categorical features among the total 15 features in our primary dataset. For categorical attributes, we used label encoding technique for data conversion from categorical to numerical ones. For example, the three values in the “Quarrel between parents” feature were changed as follows: the value allocated to “Absent” was given to 0, the value assigned to “No” to 1, and the value given to “Yes” to 2.

Then, we focused on feature selection and ranking to understand which feature is mostly responsible for poor mental well-being and which does not.

### Feature Selection and Ranking

Feature Selection and Ranking is a process that enable us to identify the valuable features in any dataset. It is used in our experiment to readily differentiate the features that play a crucial role in predicting mental health state. A univariate feature selection approach is applied for the feature selection process. The best features are chosen by utilizing the “SelectKBest” function to extract the top best features which are shown in Fig 3. Each feature is compared to the target variable to check for a statistically significant association between them. It is obvious from the figure that, “Quarrel between parents” is the prime reason leading to the poor mental health status of a student. Secondly, “Worry about social support during Covid-19” is almost the same preference for low mental well-being. Thirdly, “Disruption of daily life” has an impact on student’s mental health status. “Stable family income”, “Fear of getting infected with Covid-19”, and “Own income” are also a consequence of the impact on mental state. We have noticed that “Worry about academic delay” is also a reason for poor mental well-being. Other features also had minimal influence on mental well-being.

**Fig. 3:**
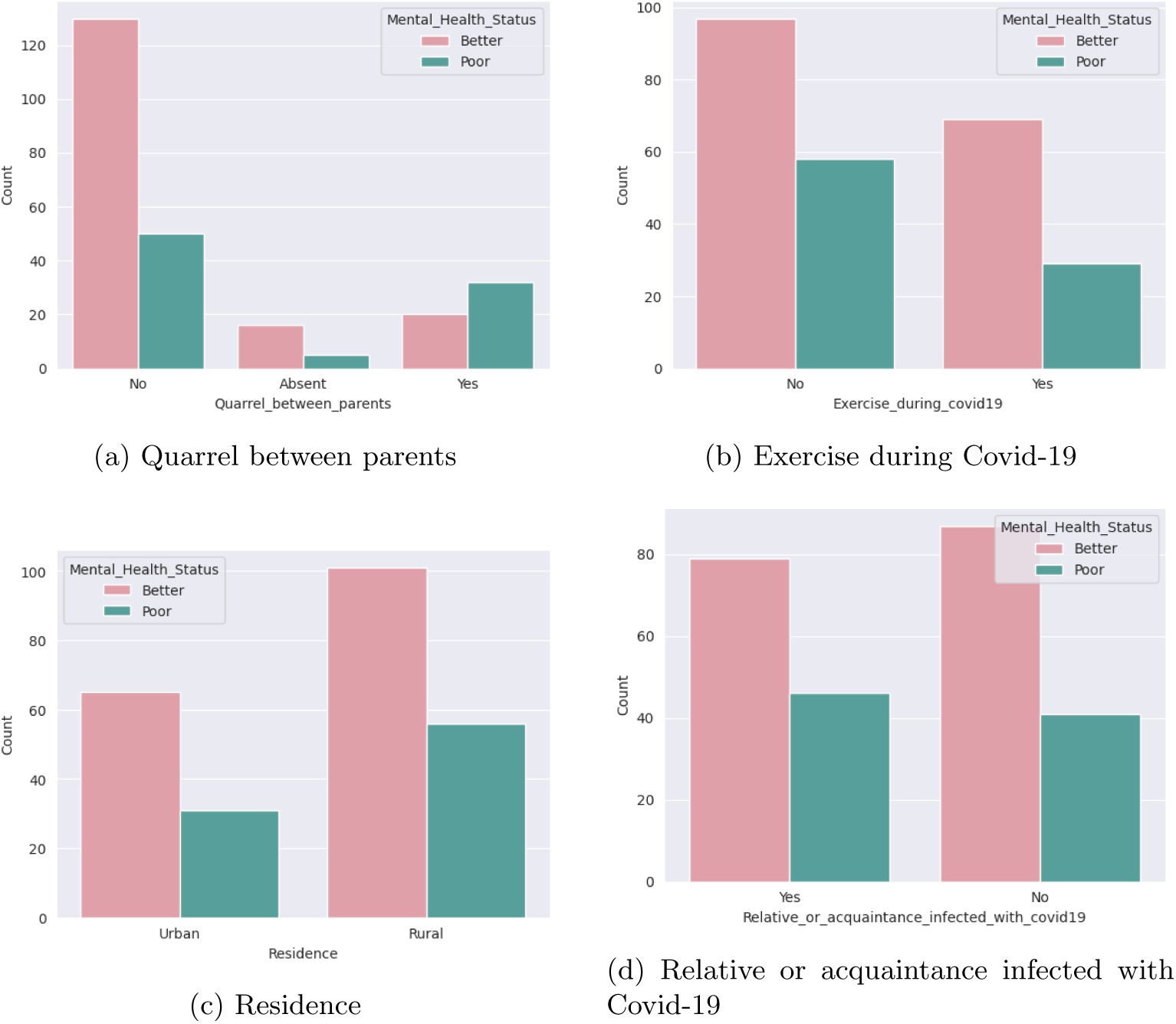
(B) Mental health status w.r.t different features (Cont.)

### Analysis of Prediction Technologies

The process of analyzing prediction technologies involves selecting from a variety of ML and DL techniques that can be utilized in the experimentation. In this case, several predictive models were used, such as those based on classification and neural network algorithms. The goal is to identify the best classifier for analyzing the problem. Each classifier must therefore be trained on the featured set, and the classifier with the best classification results is used for prediction. The classification and neural network algorithms taken into consideration are:

### Machine Learning Models

To determine the level of mental wellness, ten ML classifiers were used based on their performance. The following is a list of all the supervised ML techniques with theoretical explanation we applied in our experiment.

#### Random Forest

There are a number of decision trees that make up the RF. Input data is used to train a set of decision trees. This algorithm generates a large number of decision trees from random subsets of a dataset, and then uses the predictions from each tree to forecast the final result based on the predictions that received the most votes. With an ensemble technique that outperforms the basic tree structure [43].

#### CatBoost

Categorical Boosting is referred to as CatBoost, a new gradient-boosting method that does a better job of dealing with categorical characteristics [44]. It also uses a novel method for calculating leaf values while selecting the tree structure, which helps reduce overfitting. Consequently, this new algorithm outperforms the previous methods, such as Gradient Boosted Decision Trees (GBDT), XGB, and LGBM.

#### Extreme Gradient Boosting

XGB is a scalable ML method that applies a gradient boosting architecture for tree boosting. It is an end-to-end tree-boosting method that is commonly used in data research [45]. It can address real-world size issues with far fewer resources.

#### Support Vector Machine

The SVM is a supervised ML technique used for classification and regression tasks [46]. It’s primary goal is to find the optimal hyperplane that directly separates the information focused on two-part by augmenting the edge.

#### K-Nearest Neighbour

The KNN algorithm is a popular classification approach in ML and pattern recognition. The approach works on the idea that if the majority of the k most comparable samples to a query point q in the feature space belong to a specific category [47], then the query point q must also belong to that category.

#### Decision Tree

It is also a supervised ML strategy that constructs a classification model in a tree and produces classification rules in a top-down manner [48]. It also generates rule sequences used to determine the class of a new observation. Sometimes post-pruning approaches are employed to dealing with overfitting issues.

#### Gradient Boosting Machine

The learning mechanism in gradient boosting machines, or simply GBMs, fits new models sequentially to estimate the response variable more accurately. The basic concept behind this technique is to build new base-learners that are maximally correlated with the negative gradient of the loss function, which is connected with the entire ensemble [49]. The loss functions used can be arbitrary, however for a better understanding, if the error function is the conventional squared-error loss, the learning approach will result in successive error-fitting.

#### AdaBoost

AdaBoost is a common approach for building a robust classifier from a linear combination of member classifiers. During the training process, the member classifiers are chosen to reduce mistakes in each iteration step [50]. AdaBoost provides a straightforward and practical approach for generating ensemble classifiers.

#### Multinomial Naive Bayes

The MNB classifier is one of the NB classifier variants frequently used as a baseline in text categorization. However, this classifier is not entirely Bayesian [51].

#### Light Gradient Boosting Machine

LGBM is a free and open source distributed gradient boosting framework for ML that was created by Microsoft. It uses decision tree algorithms to do ranking, classification, and other ML tasks.

### Deep Learning Models

Despite using ML, to assess more accurately the level of mental wellness in different structures, four DL techniques were utilized, considering their performance in this experiment. The following is a is a list of the neural network techniques we applied in our experiment.

#### Convolutional Neural Networks

CNNs have been successfully used in many applications where important information about data is embedded in the form of features, such as speech and image [52]. However, CNNs offer noticeable advantages for tabular data analysis, especially in scenarios where data proves to exhibit relationship or sequential patterns in the tabular data, likewise appropriate in our dataset.

#### Recurrent Neural Networks

The RNNs are efficient in learning from past experiments and can simultaneously process a sequence of inputs and outputs, which is a kind of sequence-to-sequence network that is exceptionally efficient in handling genomic data [53]. In our cases, features in tabular data have an implicit sequence or hierarchical structure that an RNN can exploit.

#### Hybrid Neural Networks

A Hybrid Neural Network is a kind of network that combines elements from several architectures rather than being solely based on one specific architecture [54].

#### Siamese Neural Networks

SNNs, where similarity has always been a key aspect, are typically used for verification and identification tasks [55]. In this network, it first examines the features to determine their similarity. Instead of learning to classify inputs, it learns to differentiate between two inputs.

### Evaluation Metrics

To select the best performing model for the prediction of mental health status, the performance of all the classifiers was evaluated using metrics such as Accuracy, Precision, Recall, F1-score, and Area under the curve (AUC).

#### Accuracy

The accuracy metric, in general, measures the proportion of right predictions to the total number of instances examined [56]. Here, ‘TP’ stands for true positive, ‘TN’ stands for true negative, ‘FP’ stands for false positive, and ‘FN’ stands for false negative.

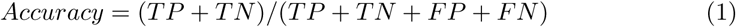

#### Precision

Precision is used to calculate the percentage of positive patterns that are accurately predicted out of all predicted patterns in a positive class [56].

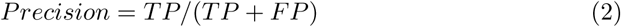

#### Recall

When we need to identify the number of positives that may fairly be predicted, recall is another viable selection of evaluation metric [57]. The recall is used to measure the fraction of positive patterns that are correctly classified.

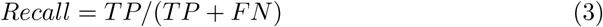

#### F1 Score

The F1 score maintains a balance between accuracy and recalls for any classifier [57]. An F1 score is a number between 0 and 1, representing the concordant accuracy and recall measures.

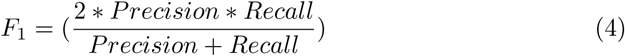

#### Area Under the Curve (AUC)

The Area under the ROC curve (AUC) indicates how well a model can discriminate across classes (the positive and negative classes). The higher the AUC, the better the model predicts 0’s as 0 and 1’s as 1 [58].

## 4 Experimental Results Analysis

In the dataset, there are 14 features and one label of which 13 features are categorical types and two are numerical ones. Before delving into the result analysis of ML and DL techniques, we conducted Chi-Square test to find the associativity of the features with respect to levels. Then we analyzed our ML and DL models’ performance in terms of Accuracy, Precision, Recall, F1-score, and AUC.

### Finding Associative Features using Chi-Square Test

Chi-Square test technique works with the target column and features by considering the important variables. Mostly it is used where the target variable is categorical, and the likelihood aspect is maintained while evaluating the statistical significance. It examines the degree of any inconsistencies between the actual and expected results based on the variables involved in the relation of the sample size and number. Degrees of Freedom (DOF) are being applied to determine whether a null hypothesis can be rejected or not based on the total number of experimental variables and samples. We have applied the Chi-Square test and found the five most associative features (“Stable family income”, “Disruption of daily life”, “Own income”, “Sleep status”, and “Fear of getting infected with Covid-19”) which was presented in Table 2. Following that, we applied ML models considering these five features only and found 72.54 accuracy while applying RF. Here, with a 95% confidence interval (CI), and a p-value of less than (or equal to) 0.05 was considered to be significant. The DOF and critical value were also mentioned in the Table 2.

**Table 2:** Analysis of associations between different features.

| Feature | Chi2 | P-value | DOF | Critical value |
| --- | --- | --- | --- | --- |
| Gender | 0.073 | 0.787 | 1 | 3.841 |
| Worry about social support during Covid-19 | 15.156 | 9.897 | 1 | 3.841 |
| Residence | 0.170 | 0.680 | 1 | 3.841 |
| Stable family income | 6.660 | **0.009** | 1 | 3.841 |
| Living with parents | 0.516 | 0.472 | 1 | 3.841 |
| Relative or acquaintance infected with Covid-19 | 0.443 | 0.505 | 1 | 3.841 |
| Worry about academic delay | 2.632 | 0.104 | 1 | 3.841 |
| Disruption of daily life | 11.886 | **0.00056** | 1 | 3.841 |
| Exercise during Covid-19 | 1.301 | 0.253 | 1 | 3.841 |
| Own income | 4.275 | **0.038** | 1 | 3.841 |
| Quarrel between parents | 21.516 | 2.126 | 2 | 5.991 |
| Sleep status hour during Covid-19 | 10.492 | **0.005** | 2 | 5.991 |
| Internet browsing hours during Covid-19 | 0.940 | 0.815 | 3 | 7.814 |
| Fear of getting infected with Covid-19 | 5.419 | **0.019** | 1 | 3.841 |

### Prediction Analysis using ML and DL

The proposed methodology of our experiment, which was already depicted earlier. The whole methodology is divided into different steps to do our research efficiently. The first step is the data collection and it’s description, which was also previously mentioned. Following data preprocessing step, we split our dataset into two subsets using the “train test split” function: training (80%) and testing (20%). We used the “stratify” parameter when splitting the dataset to keep the class levels balanced in both the training and test subsets, matching the distribution of the collected dataset [59]. Then, we trained ten ML techniques using our processed dataset. The Python programming language (Python 3) has been used to fit the dataset to ML techniques. This technique included RF, CatBoost, XGB, GBM, SVM, DT, K-NN, LGBM, AdaBoost, and MNB. After learning the insight and figuring out the pattern of the training subset, then we provided a testing subset to evaluate the model how accurately predicted on new data samples. Lastly, the performance of the classifiers was compared using performance metrics such as Confusion Matrix, Precision, Recall, F1-score, and AUC Score. The performance metrics for all the applied ML models are presented in Table 3. It is evident from this table that “accuracy” is the best evaluation metric for predicting mental health status.

**Table 3:** Result analysis over implemented ML algorithms.

| Model | Accuracy | Precision | Recall | F1-score | AUC |
| --- | --- | --- | --- | --- | --- |
| RF | **0.86** | **0.87** | **0.86** | **0.86** | **0.81** |
| CatBoost | 0.82 | 0.83 | 0.82 | 0.82 | 0.77 |
| XGBoost | 0.81 | 0.80 | 0.80 | 0.80 | 0.76 |
| GBM | 0.78 | 0.81 | 0.78 | 0.76 | 0.70 |
| SVM | 0.76 | 0.76 | 0.76 | 0.76 | 0.71 |
| LGBM | 0.76 | 0.76 | 0.76 | 0.76 | 0.71 |
| KNN | 0.74 | 0.77 | 0.75 | 0.71 | 0.65 |
| AdaBoost | 0.72 | 0.72 | 0.73 | 0.70 | 0.64 |
| DT | 0.72 | 0.72 | 0.73 | 0.72 | 0.68 |
| MNB | 0.70 | 0.73 | 0.71 | 0.65 | 0.59 |

In comparison, the RF model yielded the highest accuracy of 86 percent outperformed other models. It also had decent precision (87), recall (86), and f1-score (86) scores compared to other classifiers. The next best classifier is CatBoost which achieved performance over 80 in terms of evaluation metrics. A bar chart depicted in Fig 4 illustrates the performance of all the classifiers in which the RF surpasses other techniques. Considering AUC, again RF achieved the highest score compared to the other models, which is 81 as shown in Fig 5.

**Fig. 4:**
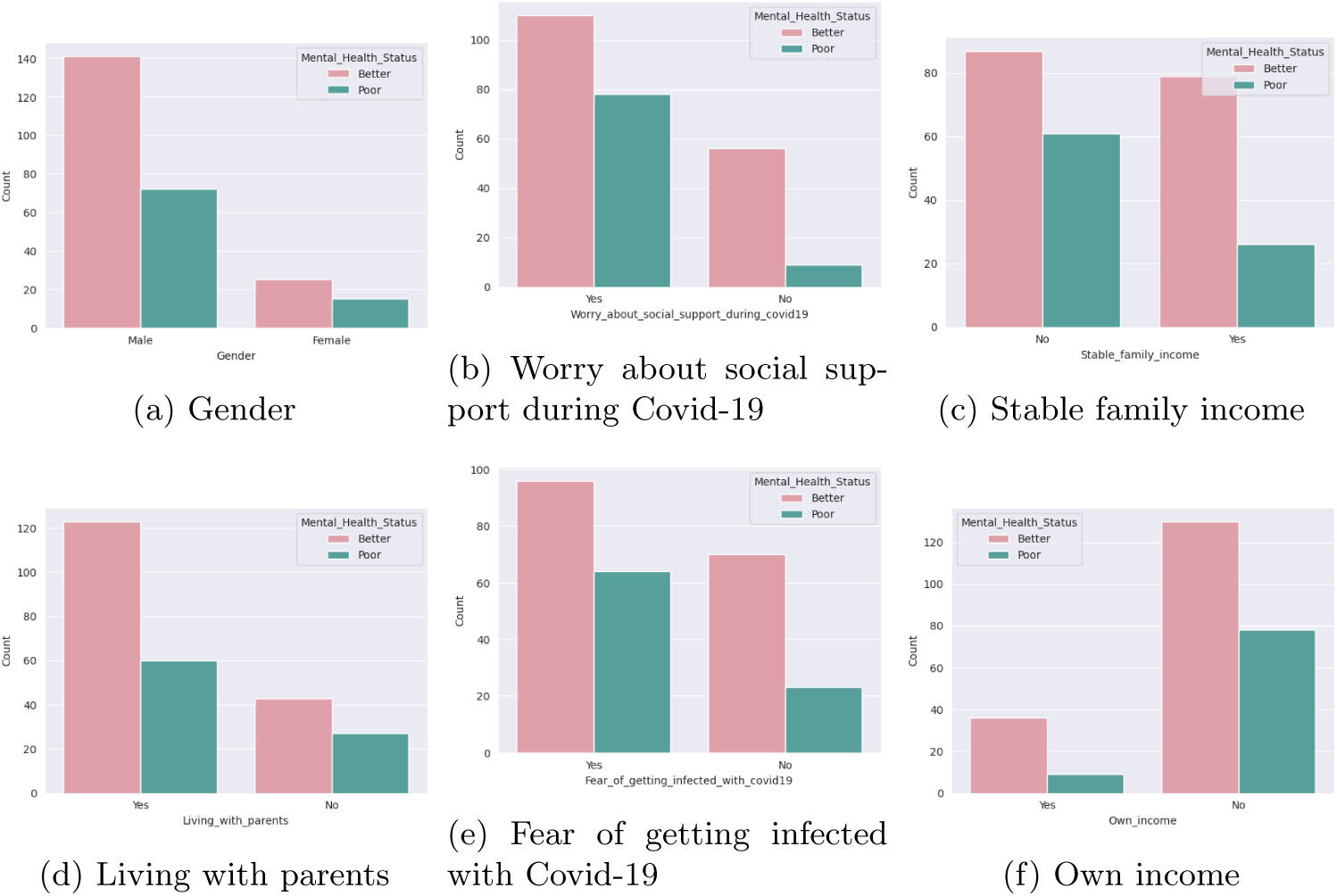
(C) Mental health status w.r.t different features.

**Fig. 5:**
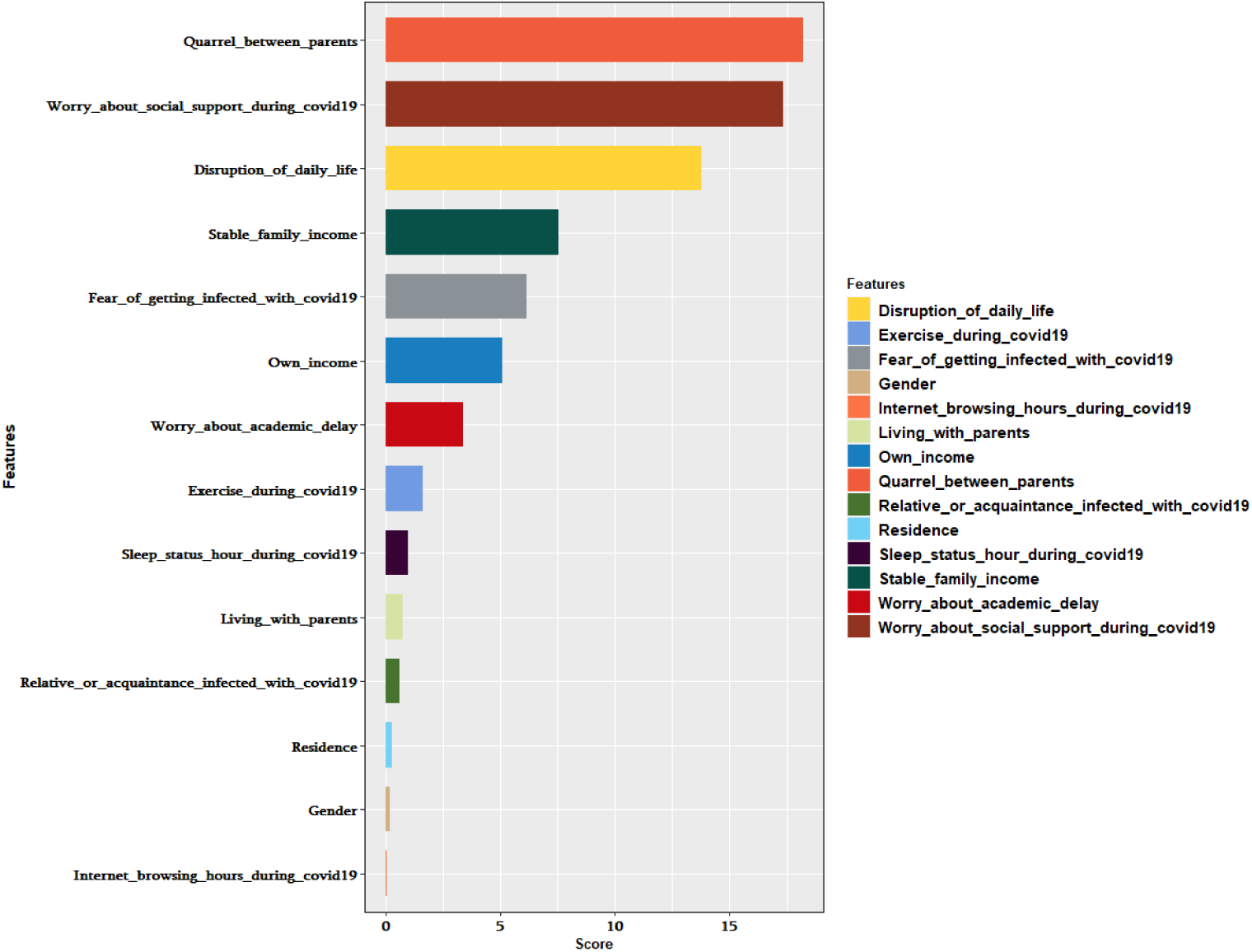
Features with scores.

In DL result analysis, we followed the same data pre-processing technique as mentioned in ML prediction analysis. Afterward, we used our processed dataset to apply four different DL methods. These methods included CNNs, RNNs, Hybrid Neural Networks, and SNNs. Once the model gained insights and identified patterns within the training subset, it then moved on to evaluate the accuracy of predicting new data samples by utilizing the testing subset. Additionally, we compared the performance of the classifier using the confusion matrix similar to what we devoted in ML. The performance metrics for all the DL classifiers are detailed in Table 4.

**Table 4:** Result analysis over implemented DL techniques.

| Model | Accuracy | Precision | Recall | F1-score | AUC |
| --- | --- | --- | --- | --- | --- |
| **SNN** | **0.75** | **0.81** | **0.70** | **0.67** | **0.72** |
| CNN | 0.72 | 0.60 | 0.57 | 0.58 | 0.70 |
| Hybrid | 0.69 | 0.56 | 0.53 | 0.54 | 0.71 |
| RNN | 0.70 | 0.61 | 0.63 | 0.64 | 0.69 |

From the table, it reveals that the SNN demonstrates the highest accuracy of 75 percent. While the CNN model also performs well, its accuracy (72%) is slightly less than SNN. SNN excels due to its ability to effectively handle common class imbalance issues and identify similar features within the dataset, requiring fewer samples per class during training. Additionally, it had the best precision (81), recall score (70), and F1-score (67) compared to other classifiers, which is presented in Table 4. In most cases, an AUC of 0.5 typically indicates no discrimination, meaning the model’s ability to differentiate between classes based on the test dataset. An AUC range between 0.7 to 0.8 is generally considered acceptable [60]. In this study, the SNN demonstrated an AUC of (72%), outperforming other models which are depicted in Fig 6.

**Fig. 6:**
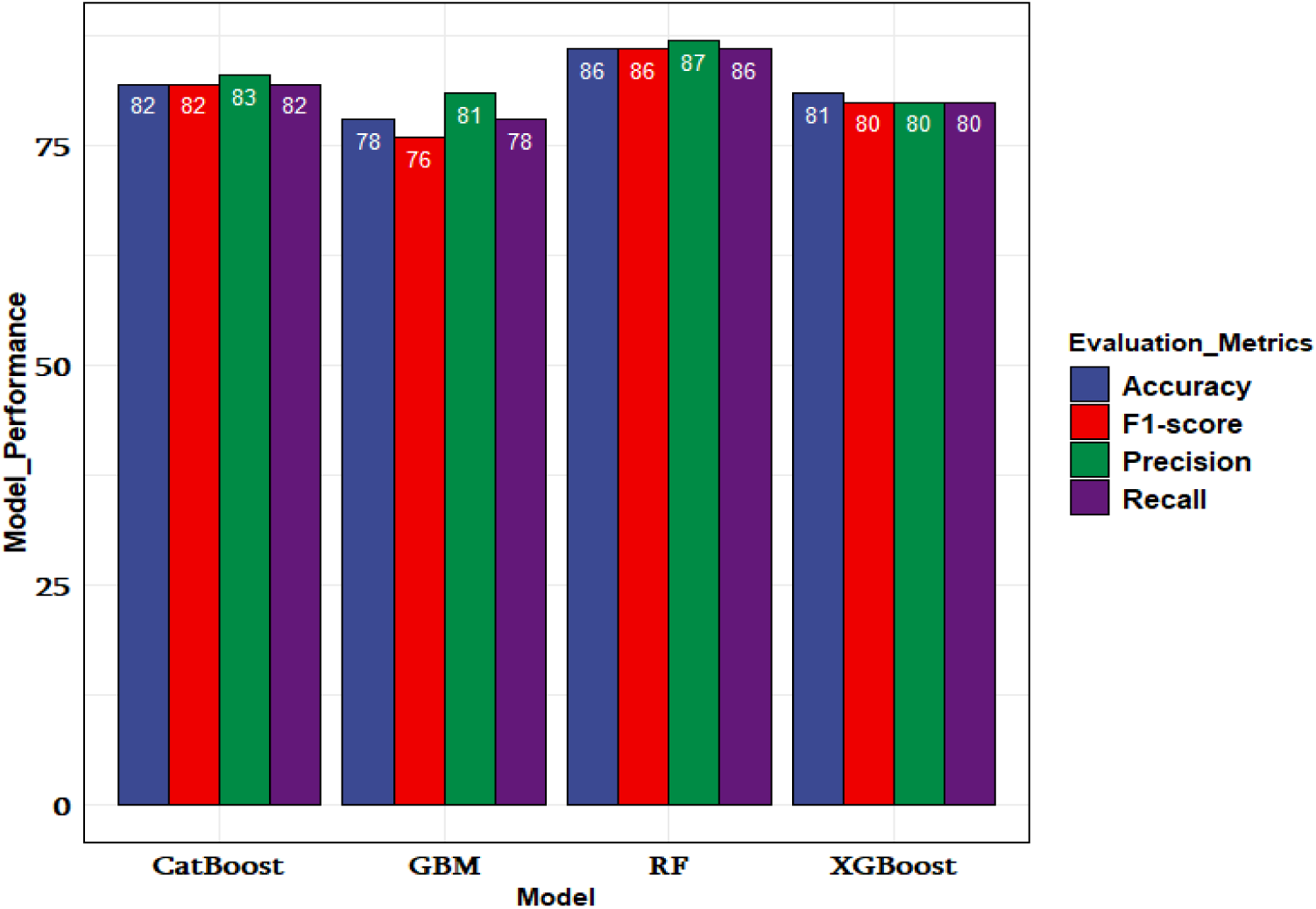
Comparing the performance of all ML models.

**Fig. 7:**
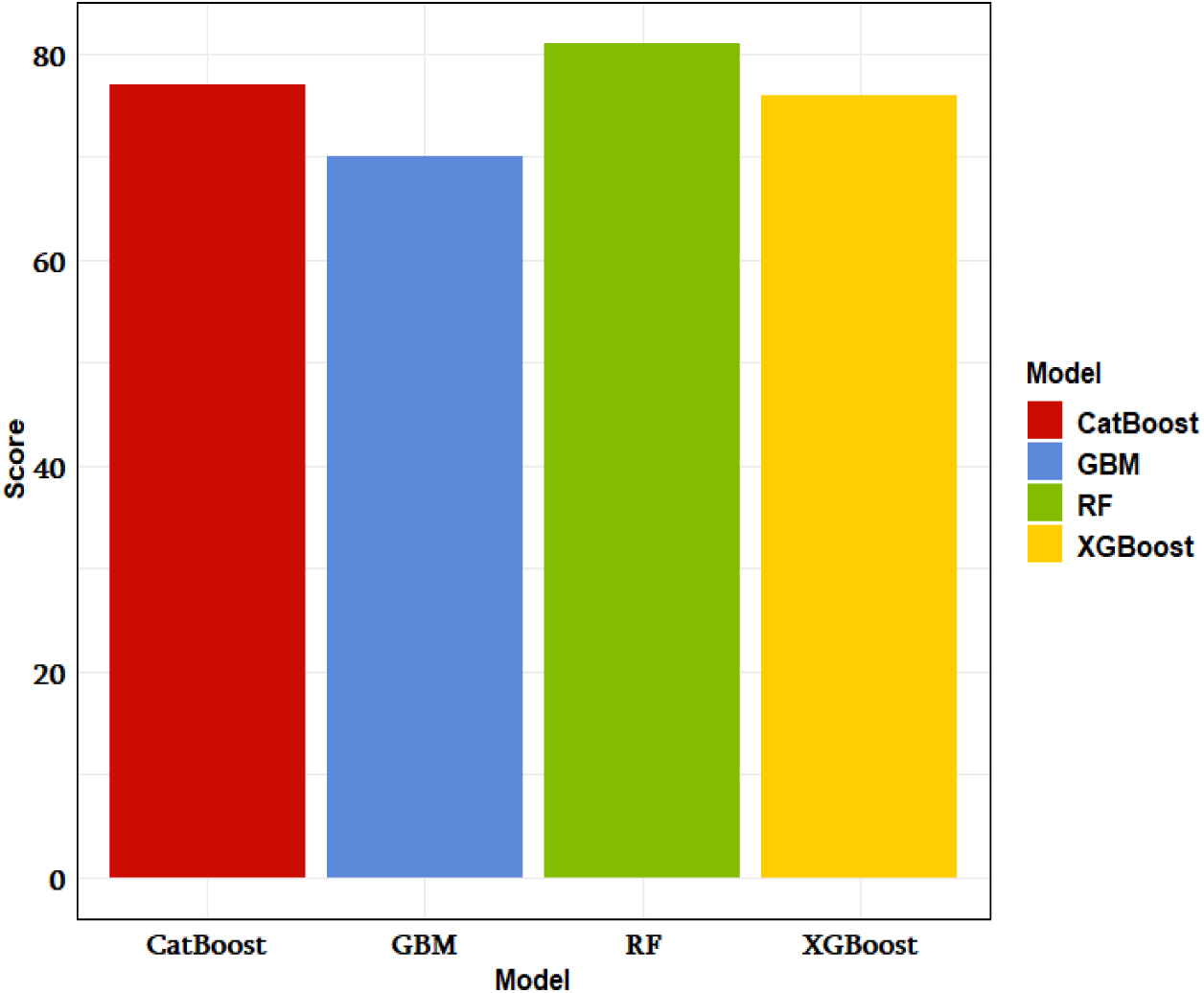
Comparing the AUC of ML models.

**Fig. 8:**
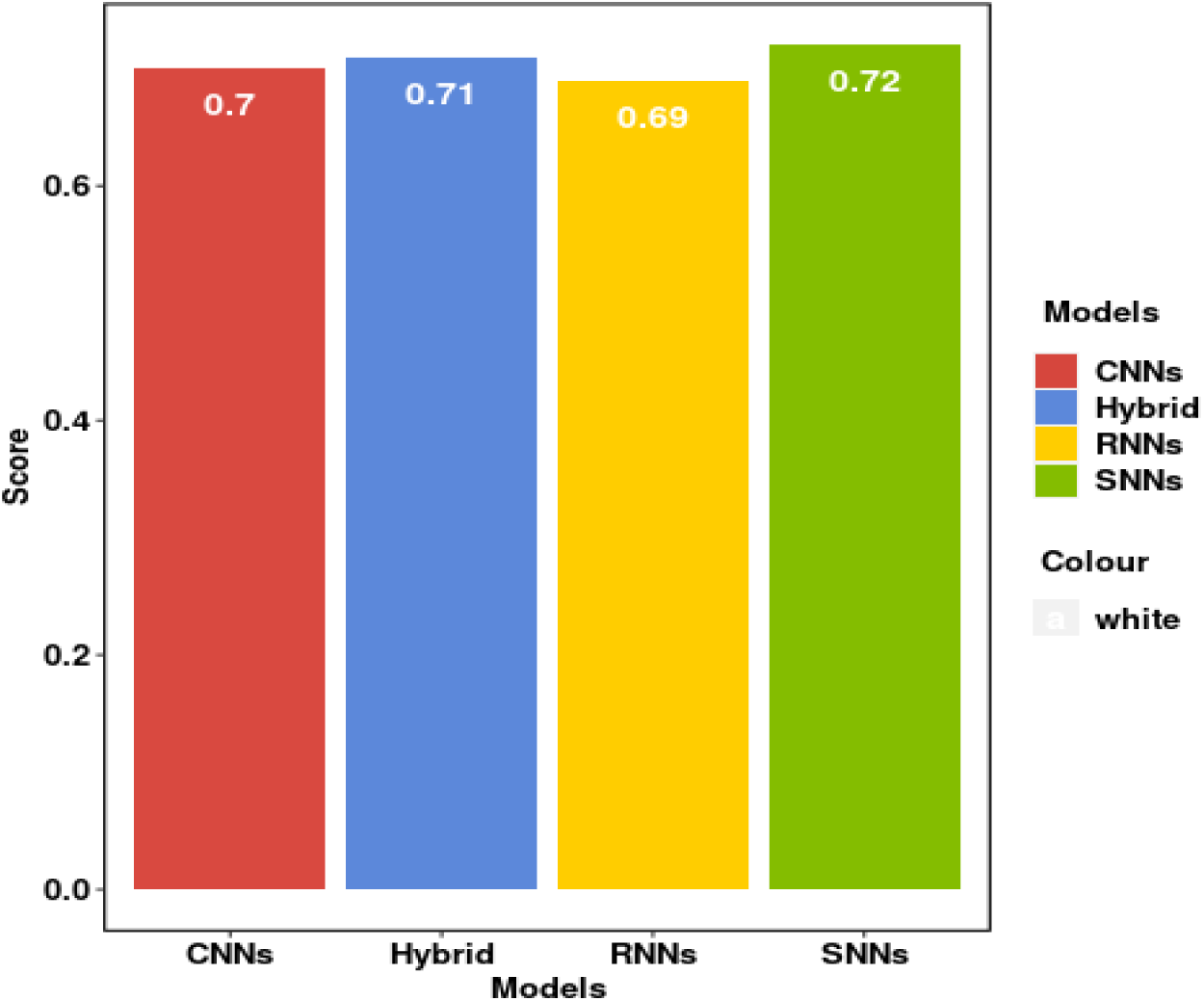
Comparing the AUC of DL models.

## 5 Discussion

The analysis of [36] pointed out that Covid-19 lockdowns had a variety of consequences, including increased psychological pressure on the daily lives of students and parents. The authors also indicated the effects of the epidemic in many areas of life, particularly in the education sector. Lack of digital skills, electricity, accessibility, facilities, and network challenges are the major obstacles that students face when enrolling in online education during this epidemic that contributed to students’ poor mental status [61]. The researchers also discovered that over 97 percent of university students were experiencing mental health issues due to the arising of the current epidemic. The primary goal of our study is to examine the psychological status of Bangladeshi university students during and after the current pandemic.

The emergence of the Covid-19 prevalence is presently posing a severe threat to world health. Bangladeshi students are not safe from this threat also. To improve university students’ mental health, it is essential to conduct a mental health assessment as soon as possible in Bangladesh. Manually doing the examination can be difficult; however, ML and DL provide a promising technique for performing the tasks in this pandemic situation. Our study conducted a Chi-Square statistical test (at p less than 0.05) to identify the five most significant features: “Stable family income”, “Disruption of daily life”, “Own income”, “Sleep status (hour) during Covid-19”, and “Fear of getting infected with Covid-19”. It shows that students from rural areas have a better mental health status than students from urban areas. This might be because rural regions have a lower Covid-19’s impact rate. We also discovered that students who are concerned about academic delays tend to have poorer mental health statuses. In our experiment, RF, CatBoost, and XGB performed better than the other classifiers during testing. Algorithmically, the RF model tackles the overfitting problem of decision trees by implementing randomization to achieve better generalization and outperform the other classifier. Because of these reasons, in this paper, it (RF) achieved the highest accuracy 86% than other classifiers. Our dataset has many categorical features, and CatBoost generally works well for categorical types of features. Therefore, in this research, we have applied this gradian boosting algorithm. On the other hand, XGB attempts to create new trees that complement the existing ones and boost training for difficult-to-classify data points. At last, to evaluate the performance of the classifiers, we use the measured area under the ROC curve, also known as AUC. In comparison, the larger the AUC, the higher the model’s performance. The RF classifier surpasses the other models by having the greatest AUC and the lowest False positive rate.

Elsewhere, we evaluated four DL algorithms where SNN shows the highest accuracy compare to the others. It outperforms when the features are alike to each other. Additionally, this method is good at handling common class imbalance issues and finding similar features in the dataset. It also worked well in our study while needing fewer samples for training each class. Our second best-performing DL model was CNN and it performs well where data contains categorical relationships.

### Strengths and Limitations

Strengths in this research, include 1) this research was conducted during the pandemic, focusing particularly on university students in Bangladesh, where mental health research is limited; 2) we collected a dataset from diverse dimensions such as 16 universities; 3) we also achieved decent enough accuracy by applying well-known classification models to predict accurately whether a student’s mental health status is poor or better. Along with some substantial advantages, there are some limitations in this experiment. Limitations are 1) we cannot include all the university students’ data, hence the findings of our analysis may not be generalizable to all Bangladeshi students; 2) as the data is self-reported via an online questionnaire, some of it may be biased; 3) the dataset was limited to build more robust prediction; additional data could be added to improve forecast accuracy.

## 6 Conclusion

Globally, Covid-19 has impacted negatively on people’s lives, and it has hampered individuals’ physical and mental well-being, especially university students in Bangladesh. In this paper, we employed AI experiments and statistical test on our primary dataset where data was collected through an online questionnaire. Then we trained the dataset with ML and DL techniques where RF and SNNs revealed the best accuracy in predicting mental health state. On the other hand, in the Chi-Square feature test, we found the five most associative and significant predictors that greatly influence the psychological health conditions of students during the pandemic in Bangladesh. We hope that our findings will assist university administrators and healthcare providers in maintaining the mental health of students during and aftermath the epidemic. Based on our findings, further research is needed to analyze the long-term impact of the Covid-19 pandemic on students’ mental health performance. In the future, we would add more features to the dataset to track the development of student’s mental and physical status over a more extended period, such as several years. As our dataset size is limited, more students’ records from educational institutions could be included in future studies to get more reliable and robust predictions.

## Data Availability

Based on request data set will be shared.

